# Transcutaneous auricular vagus nerve stimulation regulates subjective fear in naturalistic contexts via modulation of prefrontal neural dynamics

**DOI:** 10.64898/2026.08.07.26359962

**Authors:** Can Liu, Kun Fu, Qi Liu, Xiaodong Zhang, Siyu Zhu, Xinqi Zhou, Rong Zhang, Benjamin Becker, Keith M. Kendrick, Weihua Zhao

**Affiliations:** The Center of Psychosomatic Medicine, Sichuan Provincial Center for Mental Health, Sichuan Provincial People’s Hospital, University of Electronic Science and Technology of China, Chengdu, 611731, China; Peking Union Medical College, Chinese Academy of Medical Sciences, Beijing, China; Research Unit of NeuroInformation, Chinese Academy of Medical Sciences, Chengdu, China; School of Sport Training, Chengdu Sport University, Chengdu, 610041, China; Institute of Brain and Psychological Sciences, Sichuan Normal University, Chengdu, 610066, China; Neuroscience Research Institute; Key Laboratory for Neuroscience, Ministry of Education of China; Key Laboratory for Neuroscience, National Committee of Health and Family Planning of China; and Department of Neurobiology, School of Basic Medical Sciences, Peking University, Beijing, 100191, China; The Department of Psychology, The University of Hong Kong, Hong Kong, 999077, China; China-Cuba Belt and Road Joint Laboratory on Neurotechnology and Brain-Apparatus Communication, University of Electronic Science and Technology of China, Chengdu, 610054, China

**Keywords:** Transcutaneous auricular vagus nerve stimulation, continuous fear evaluation, naturalistic paradigm, fNIRS, spatial activation patterns, anxiety

## Abstract

Although non-invasive transcutaneous auricular vagus nerve stimulation (taVNS) has demonstrated a therapeutic-relevant potential by enhancing mood recovery and fear extinction, its influence on neural dynamics during naturalistic, sustained fear processing remains unclear. In this study, we employed a randomized, sham-controlled, parallel-group design involving 63 participants (taVNS: n = 33; sham: n = 30) who provided continuous subjective fear ratings (1170 timepoints) while watching a 10-minute fear-inducing video, with simultaneous fNIRS recordings. We employed: (1) a convolutional neural network (CNN) to decode fear ratings from frontal activations, (2) validation of stimulus-evoked activity comparing fNIRS with fMRI signal, (3) dynamic conditional correlation analysis to assess taVNS-induced connectivity changes, and (4) moderation analysis to examine anxiety state effects. Behaviorally, taVNS significantly attenuated fear responses during four threat phases by content analysis: T1 (ghost appearance), T2 (escape sequence), T3 (sudden threat emergence) and T4 (suicide scene). Neurally, taVNS suppressed medial prefrontal cortex (mPFC) activation during escape (T2) and disrupted the typical fear coupling between fear experience and brain activity. Furthermore, taVNS enhanced intra-mPFC functional connectivity, suggesting a potential neural basis for modulating subjective threat appraisal. Additionally, state anxiety significantly moderated brain-behavior relationships. These findings demonstrate that taVNS attenuates fear responses through modulation of mPFC engagement and strengthening frontal network integration. Our results highlight taVNS as a promising neuromodulatory intervention for fear-related disorders (e.g., anxiety disorder), particularly as an early adjunct to exposure-based therapies, warranting further clinical validation.

## Introduction

Fear serves as an evolutionarily conserved adaptive response that protects against environmental threats [1]. However, when dysregulated, excessive fear responses can become maladaptive, contributing to anxiety disorders [2], post-traumatic stress disorder (PTSD) [3] and other fear-related psychopathologies [4]. These conditions are characterized by persistent, exaggerated threat processing, heightened subjective fear and impaired fear extinction, a key mechanism in exposure-based therapies [1, 5]. Given that mental disorders are defined by their subjective experiences, it is unsurprising that treatments neglecting these subjective mental qualities have shown limited efficacy [6]. Due to the shortcomings of current therapies—such as partial effectiveness and high relapse rates—there is an urgent need for novel interventions targeting the neural substrates of subjective fear.

The prefrontal cortex (PFC) serves as a pivotal hub for fear processing through its dual functions of integrating sensory and emotional signals [7] and subjective fear modulation [8]. Notably, specific prefrontal subregions, particularly the medial prefrontal cortex (mPFC), are critically involved in fear regulation, a fundamental adaptive mechanism that enables behavioral flexibility and social cohesion [9, 10]. The mPFC executes this regulatory function through two key mechanisms: (1) facilitating fear extinction learning via governing subcortical systems and (2) actively suppressing fear responses during subsequent threat encounters [11, 12]. Additionally, mPFC also regulates autonomic arousal [13], however on the neural level the autonomic arousal and subjective fear/anxiety/arousal experience are different [14–16]. Emerging evidence suggested that transcutaneous auricular vagus nerve stimulation(taVNS) may alter prefrontal activity during emotional inhibition [17] and enhance the dynamic interplay between interoceptive awareness and cognitive processing by modifying functional connectivity between the insula and prefrontal regions [18].

A growing number of studies suggests taVNS, a novel non-invasive neuromodulation technique targeting the auricular branch’s cutaneous receptive field [19], may effectively modulate pathological fear responses. By activating vagal afferents, taVNS influences central and peripheral neural circuits, including the nucleus of the solitary tract (NTS), the locus coeruleus-norepinephrine (LC-NE) system, hippocampus, amygdala and prefrontal cortex [19–21].Clinically, taVNS has demonstrated therapeutic potential across multiple domains: reducing anxiety symptoms [22, 23], alleviating depression [24–26], enhancing cognitive performance [27] and improving sleep in chronic insomnia [28]. Furthermore, taVNS appears to facilitate emotional processing through improved emotion recognition [29, 30] and enhanced inhibitory control [17], while also promoting fear extinction [31] and reducing fear or fear-related behavior in both laboratory animal and human research [31–34], the core inhibitory process underlying exposure therapy, likely modulated via noradrenergic system activation [35, 36]. While subjective fear is a primary contributor to anxiety disorders, its neural basis differs from that of a conditioned, unextinguished threat [37, 38], it remains unclear whether its laboratory-observed neural mechanisms translate to more dynamic real-life contexts where fear naturally emerges [39], despite one study reporting that taVNS therapy behaviorally improved PTSD symptoms in a naturalistic condition [35].

While traditional fear-conditioning paradigms (i.e. Pavlovian conditioning) have provided foundational insights, naturalistic fear paradigms (e.g., dynamic movie-viewing tasks) offer distinct advantages for studying neural processing of threat-related sensory information [39, 40] and can facilitate testing of novel interventions for fear in dynamic and close to real-life contexts [41]. These ecologically valid approaches preserve the temporal dynamics of brain activity during continuous audiovisual stimulation, maintaining a direct correspondence with real-world perceptual experiences while allowing investigation of integrated multisensory processing [40]. Recent studies indicate that higher-order regions, including the medial prefrontal cortex (mPFC), play a critical role in processing affective experiences during naturalistic stimuli, with distinct neural substrates linked to different emotional expressions [42]. Despite these advances, the neuromodulatory effects of taVNS on prefrontal functioning during naturalistic fear evaluation remain poorly understood, representing a critical gap in our knowledge of potential therapeutic applications. Detailed, mechanistic studies are needed to clarify how taVNS modulates prefrontal regions during naturalistic fear processing.

Functional near-infrared spectroscopy (fNIRS) is an increasingly utilized optical neuroimaging technique that measures hemodynamic responses (e.g., oxygenated hemoglobin; HbO) with higher temporal resolution and greater tolerance to motion artifacts compared to other methods[43, 44]. These advantages make it particularly suitable for naturalistic video-watching paradigms combined with continuous subjective evaluation. Additionally, fNIRS allows a flexible setup, enabling concurrent taVNS administration either on the earlobe (sham condition) or tragus (active condition). Thus, in the current study, we employed a pre-registered sham-controlled, participant-blinded, between-subjects design to investigate the neuromodulatory effects of taVNS on response to 10-minute video-induced fear, integrating continuous fear ratings (i.e. 1170 ratings) with fNIRS-based prefrontal cortex (PFC) monitoring. We aimed to (1) validate fNIRS-derived hemodynamic changes in the PFC through a parallel fMRI study with the same video stimulus; (2) determine whether prefrontal activity can predict continuous fear ratings using a convolutional neural network (CNN) model; (3) explore taVNS-induced alterations in behavioral fear ratings and associated neural changes and (4) examine how fear-related anxiety states moderate these associations.

## Methods and Materials

### Participants

A total of 70 right-handed healthy Chinese university students were recruited in this study. All participants had no reported history of medical or psychiatric illness, no record of long-term or regular medication use and had normal or corrected-to-normal vision. Participants were instructed to abstain from alcohol, caffeine, and nicotine on the experimental day. Participants were randomly assigned to either active taVNS or sham stimulation condition. The required sample size was estimated using G*Power (v3.1.9.7) with an a priori power analysis for a two-tailed independent-samples t test (difference between two independent means, two groups). Assuming an effect size of Cohen’s d = 0.75, an alpha level of 0.05, and 80% power with equal allocation between groups, the analysis indicated that 58 participants were required (29 per group; actual power = 0.801). Seven participants were excluded due to technical issues during fNIRS recording or collection of behavioral ratings, yielding a final sample of 63 participants (31 females, mean age 21.51 ± 2.32 years) included for further analysis (taVNS: n =33, sham: n =30). Each participant provided written informed consent and the study protocol was approved by the local ethics committee of the University of Electronic Science and Technology of China in accordance with the latest revision of the Declaration of Helsinki and was pre-registered as a clinical trial (NCT06975124, clinicaltrials.gov).

### Experimental procedure

This study utilized a sham-stimulus-controlled, between-subjects design with blinding of both participants and data analyzers. Prior to the start of the experiment, all participants were required to complete a series of mood questionnaires including the Cognitive Emotion Regulation Questionnaire (CERQ), State-Trait Anxiety Inventory (STAI), Social Interaction Anxiety Scale (SIAS) and Beck’s Depression Inventory-II (BDI-II) to control for potential confounding effects. In addition, two Positive and Negative Affect Scale (PANAS) assessments [45] and two blood pressure and pulse measurements (pre- and post-stimulation) were conducted to monitor emotional and physiological changes.

Given that the auricular branch of vagus nerve (VN) is related to touch sensation, stimulus intensity was individualized to a level above the detection threshold but below discomfort to ensure VN activation [46]. An identical calibration procedure was used for both the taVNS and sham groups (see Supplemental Methods). All participants reported being unable to distinguish whether they received active or sham stimulation. Furthermore, the researchers conducting the data analysis were kept fully blinded to group assignments throughout the study, ensuring the objectivity of both the experimental process and the results. After familiarization with the instructions of the fear rating task, participants received 15 minutes of electrical stimulation (width, 500 μs; frequency, 25 Hz, 30s on, 30s off). Finally, participants received a further 10 minutes of stimulation while completing the fear rating task, during which brain activity and skin conductance were recorded simultaneously. In the fear rating task (**Figure 1A**), participants were required to watch a 10-minute (600s) horror movie while continuously rating their subjective fear intensity from 0 to 100 (0: no fear, 100: extreme fear) in real time via mouse movements. The ratings were recorded every 0.51 seconds, yielding a total of 1170 data points. The task program was presented via Psychopy-2022.1.0.

**Figure 1.**
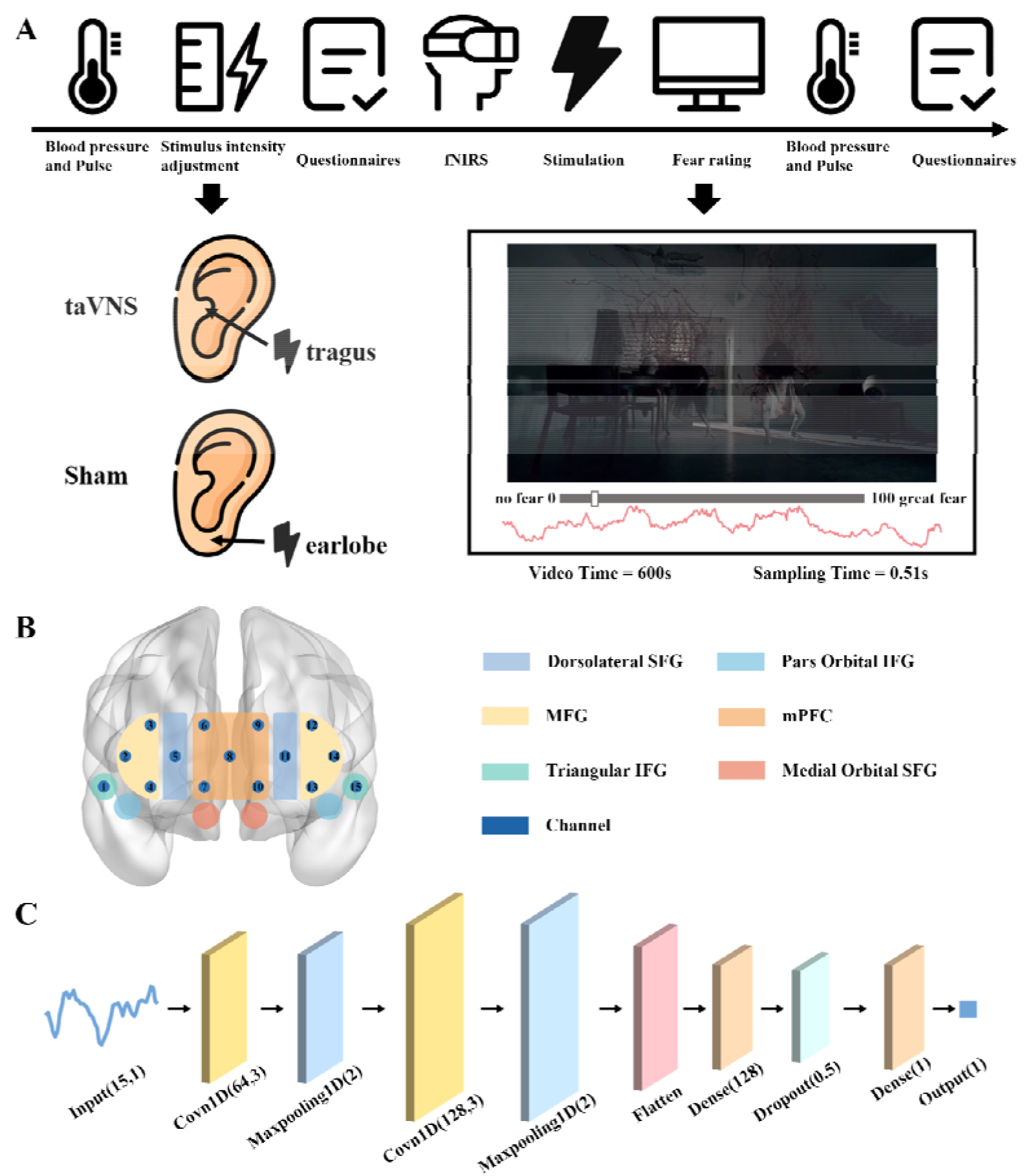
Schematic illustration of experimental protocol and data analysis. A. Experimental protocol. B. Location of fNIRS optodes over the 6 ROIs. C. Overview of convolutional neural network (CNN) analysis. SFG, superior frontal gyrus; MFG, middle frontal gyrus; IFG, inferior frontal gyrus; mPFC, medial prefrontal cortex.

At the end of the experiment, participants were required to report stimulation side effects via a seven-point Likert scale (1: not at all, 7: very much), assessing headache, nausea, skin irritation under the electrode, vigilance, unpleasant feelings, dizziness, neck pain, muscle contractions in the neck, and stinging sensation in the left ear. The full experimental procedure is detailed in **Figure 1A**.

### Transcutaneous auricular vagus nerve stimulation

The taVNS group received stimulation via a transcutaneous auricular vagus nerve stimulation device (Suzhou Huize Medical Technology Co., Ltd Model: ZR-tVNS-301, China), with ear clip electrodes attached to the left cymba conchae to target vagus nerve branches. The sham group employed electrode placement on the left earlobe, a non-vagus-innervated site (**Figure 1A**) [17]. Mean stimulation intensity was 1.20 mA (range: 0.9-1.6 mA) for taVNS and 1.17 mA (range: 0.5-1.6 mA) for sham stimulation, with no significant group difference (t = 0.498, p = 0.62).

### fNIRS data collection and preprocessing

Hemodynamic data from PFC regions were acquired using the NIRSIT LITE system (OBELAB Inc, Seoul, Korea) with LED light sources (sampling rate: 8.1367Hz, wavelengths: 760nm and 850nm). The Montag configuration included five sources and seven detectors, yielding 15 measurement channels. Channel-specific Montreal Neurological Institute (MNI) coordinates and brain region mapping are detailed in **Tables S1 and S2**. Data preprocessing was performed via the NIRS-KIT toolbox [47] in MATLAB 2023b in the following steps: (1) conversion of the raw light intensity signals to oxygenated hemoglobin (HbO) and deoxyhemoglobin (HbR) concentrations based on the Modified Beer-Lambert Law; (2) linear trend removal using a polynomial regression model; (3) motion artifacts correction with Time Derivative Distribution Repair (TDDR) algorithm [48]; (4) bandpass filtering (0.01-0.08 Hz) using a 500^th^-order finite impulse response (FIR) filter [49]. This study only focused on the HbO signal due to their superior signal-to-noise ratio in fNIRS [50].To account for hemodynamic response latency, fNIRS time series were temporally aligned with video stimuli by applying a 6-s delay correction based on the hemodynamic response function (HRF) [51].

### fMRI data validation and data analysis

Task-based fMRI data were acquired from an independent sample of 34 healthy male participants (mean age 20.79 ± 1.87 years) who viewed the same fear stimulus without real-time ratings. At the end of the task, participants were required to rate fear feelings from 1 to 9 (1, not at all; 9, very strong). Imaging data was performed on a 3.0-T GE Discovery MR750 scanner (repetition time, TR = 2 s) and preprocessed using the standard workflow in fMRIPrep 21.0.0 [52], a process based on Nipype 1.6.1. For more details see Supplemental Methods [53]. The raw temporal data was also shifted backward 6 s account for hemodynamic response delay [54].

### Validation of stimulus-evoked activity between fNIRS and fMRI

We assessed cross-modal validation of stimulus-evoked activity by comparing fNIRS (sham group) and blood oxygenation-level dependent (BOLD) fMRI signals during fear video viewing. fMRI data were processed by: (1) extracting voxels from six prefrontal ROIs (Automated Anatomical Labeling, AAL-90 atlas: dorsolateral superior frontal gyrus (SFG), middle frontal gyrus (MFG), triangular inferior frontal gyrus (IFG), par orbital IFG, medial prefrontal cortex (mPFC), medial orbital SFG, **Figure 1B**), which were overlapped with the brain regions measured by fNIRS; (2) spatially averaging voxels to obtain time-series signals within each ROI; (3) averaging across 34 participants to generate group-level time series for all 300 time points (2s for each). fNIRS signals were downsampled to 300 time points to match fMRI temporal resolution and underwent identical spatial and temporal averaging. The absence of significant sex differences in fNIRS-derived fear ratings (t = 0.14, p = 0.89) and regional activity (all |t| ≤ 1.61, ps ≥ 0.12) suggests that this sex factor is unlikely to limit the generalizability of the observed cross-modal correspondence to the fMRI dataset. Subsequently, spatial coherence was quantified using Pearson correlation coefficients between fNIRS and fMRI signals (i.e. 300 time points) for each ROI, with FDR correction for multiple comparisons (q < 0.05). To further establish the regional specificity of the fNIRS-derived signals, we conducted an additional analysis. Given the high inter-correlation among the six prefrontal fNIRS ROIs (dorsolateral SFG, MFG, triangular IFG, par orbital IFG, mPFC, and medial orbital SFG; all r ≥ 0.73, p < 0.001; **Figure S1A**), we correlated the fNIRS signals from these ROIs with the fMRI BOLD signals from four major whole-brain lobes (frontal, parietal, occipital, and temporal) to determine whether the observed coherence was specific to the frontal lobe.

### Convolutional neural network architecture

To investigate the role of PFC in fear evaluation, we employed a convolutional neural network (CNN) model (**Figure 1C**) to predict fear ratings using prefrontal activation patterns from 15 channels as input features, and used R-square (R^2^) and mean absolute error (MAE) to assess the predictive validity of the model. This CNN model incorporates two convolutional layers, each configured with 64 and 128 convolutional kernels for feature extraction. Both kernel sizes are set to 3 with a stride of 1, and rectified linear unit (ReLU) is employed as the activation function to achieve nonlinear feature transformation. Each convolutional layer is followed by a max-pooling layer, which downsamples the feature maps by retaining their maximum values. This approach highlights core salient features while effectively improving computational efficiency. The pooling kernel size and stride are set to 2 and 2, respectively [55]. Subsequently, the network flattened the extracted multidimensional features into one-dimensional vectors and fed them into a fully connected layer (dense layer) incorporating dropout regularization. This layer produced 128 output neurons, with a dropout rate of 0.5 to suppress model overfitting [56]. Finally, a linear activation function generated continuous fear scores, enabling prediction of fear levels across groups. The learning rate for model training was set to 0.001. During each training iteration, the dataset was pooled across all participants and globally shuffled, followed by random partitioning of the data, with 80% allocated to the training set and 20% to the test set.

To determine the optimal training epoch count, 10 CNN models were independently trained at each epoch within the 100–300 epoch range (step size 20). Changes in R² and MAE for both the training and test sets were dynamically monitored across different epochs (results shown in **Figure S2**). Experimental results demonstrate that model performance steadily improves with increasing epochs—both training and testing sets exhibit gradually rising R² values and consistently decreasing MAE, with consistent trends across both sets and no signs of overfitting, indicating robust model results. Beyond 200 epochs, the rate of performance improvement slows. Based on these findings, this study ultimately determined 200 as the optimal training epoch. At this parameter, 100 independent cross-validations were conducted to validate the model’s stability. This model was constructed in a Python 3.10 environment and implemented using the TensorFlow 2.15.0 framework.

### Differences in fear ratings between stimulation groups

Fear rating differences between stimulation groups were analyzed using independent samples t-tests across all 1,170 sampled time points (homogeneity of variance using Levene’s test was conducted). Key temporal windows of significance were identified through three criteria: (1) one-tailed p < 0.05 based on our priori directional hypothesis [31–34], (2) consistent effect directionality, (3) at least 10 consecutive time points (minimum 5-second duration) to distinguish sustained effects from transient noise and to support subsequent content encoding, and (4) confirmation using permutation-based cluster tests to correct for multiple comparisons. To further elucidate taVNS modulation of fear responses, we extracted video segments corresponding to significant time windows (i.e. T1-T4) and performed AI-based moonshot-v1 content analysis to characterize stimulus features associated with stimulation effects. We also conducted content analyses for additional periods with high and moderate fear ratings (1:20–1:37, 1:50–1:57, 2:55–3:02, 4:24–4:34, 5:52–5:58, 6:09–6:15, 6:30–6:38, 8:40–8:48). Furthermore, we recruited an independent sample of participants (n = 20, mean age = 24.15 ± 2.13, 10 females) to rate fear intensity for the video stimuli. We then re-analyzed the t-test between taVNS and sham groups, adding these fear ratings as a covariate. This provides quantitative evidence that stimulus fear content or fear intensity contribute to the observed localized effects of taVNS.

### Neural prefrontal activation differences between stimulation groups

To temporally align neural signals with behavioral data, we downsampled fNIRS signals to 1,170 time points, matching the temporal resolution of fear ratings. To identify significant activation differences within the four distinct fear response windows (T1-T4), we employed a permutation test with predefined cluster-based correction for each channel. The procedure involved: (1) calculating t-values via two-sample t-tests at each time point and summing them to obtain the cluster statistic t_sum; (2) identifying the cluster with the maximum t_sum and performing 10,000 permutations to generate an empirical null distribution; (3) comparing the observed t_sum against this null distribution, where clusters in the top 5% were considered statistically significant (p < 0.05). To quantify the taVNS effect, we averaged the HbO concentrations across the time windows exhibiting significant differences for each channel. Group comparisons were then conducted using independent samples t-tests with significance determined by 10,000 permutation tests. Additionally, we examined Pearson correlations between brain activity and fear ratings during significant naturalistic fear evaluation across stimulation groups. Finally, we tested whether state anxiety moderated this relationship using moderation analysis with PROCESS v3.5 (SPSS 26.0).

### Channel-based dynamic functional connectivity differences between stimulation groups

Using the significant channel identified in our activation analysis as the seed, we computed dynamic functional connectivity (dFC) matrices between the seed and all other channels for each participant. Connectivity was estimated at each time point (temporal resolution = 0.51 s) using the dynamic conditional correlation (DCC) algorithm [57], which captures time-varying or instantaneous coupling between neural signals. For the four distinct fear response windows (T1-T4), we identified time periods with significant differences in functional connectivity for each channel pair using permutation tests with predefined cluster-based correction, following the same pipeline as the activation analysis. To quantify the taVNS effect, we extracted and averaged connectivity strength within the significant time windows for each channel pair. Group comparisons were then conducted using independent samples t-tests with significance determined by 10,000 permutation tests. We evaluated Pearson correlations between dynamic functional connectivity and fear ratings during significant naturalistic fear evaluation across stimulation groups. In addition, we also performed moderation analyses using PROCESS v3.5 (SPSS 26.0) to test whether state anxiety modulated the relationship between dFC and fear ratings.

### Electrodermal activity recording and analysis

Skin conductance response (SCR) data were collected using an MP-150 BIOPAC system (BIOPAC Systems, Goleta, CA), with Ag/AgCl electrodes placed on the second phalangeal segments of the non-dominant hand’s (i.e. left hand) index and middle fingers to minimize voluntary movement artifacts during task. The raw signals digitally recorded at a sampling rate of 1000 Hz. Following the data quality check [58], six participants (three per group) were excluded for the SCR further analysis. Preprocessing involved: (1) applying a 0.05 Hz Butterworth high-pass filter to remove low-frequency noise and baseline drifts, followed by (2) downsampling to 200 Hz in MATLAB 2023b (preserving physiological signal characteristics) for analysis in Ledalab (v3.4.9) [59]. Using continuous decomposition analysis (CDA), we extracted SCR signals exceeding the 0.01 μS response threshold during horror movie viewing [60]. SCR frequency (number of responses surpassing threshold) within each significant time window served as the quantitative measure of electrodermal activity in two groups.

## Results

### Demographics and potential confounders across stimulation groups

As shown in **Table 1** and **Table S3**, the sham and taVNS groups demonstrated comparable baseline characteristics, with no statistically significant differences in demographic factors (age: p = 0.47; gender distribution: p = 0.90), emotional state (PANAS positive/negative affect scores: all p > 0.15), physiological measures (systolic/diastolic blood pressure, pulse rate: ps > 0.33), personality traits (ps > 0.11), subjective adverse reaction ratings (ps > 0.07, uncorrected).

**Table 1.**
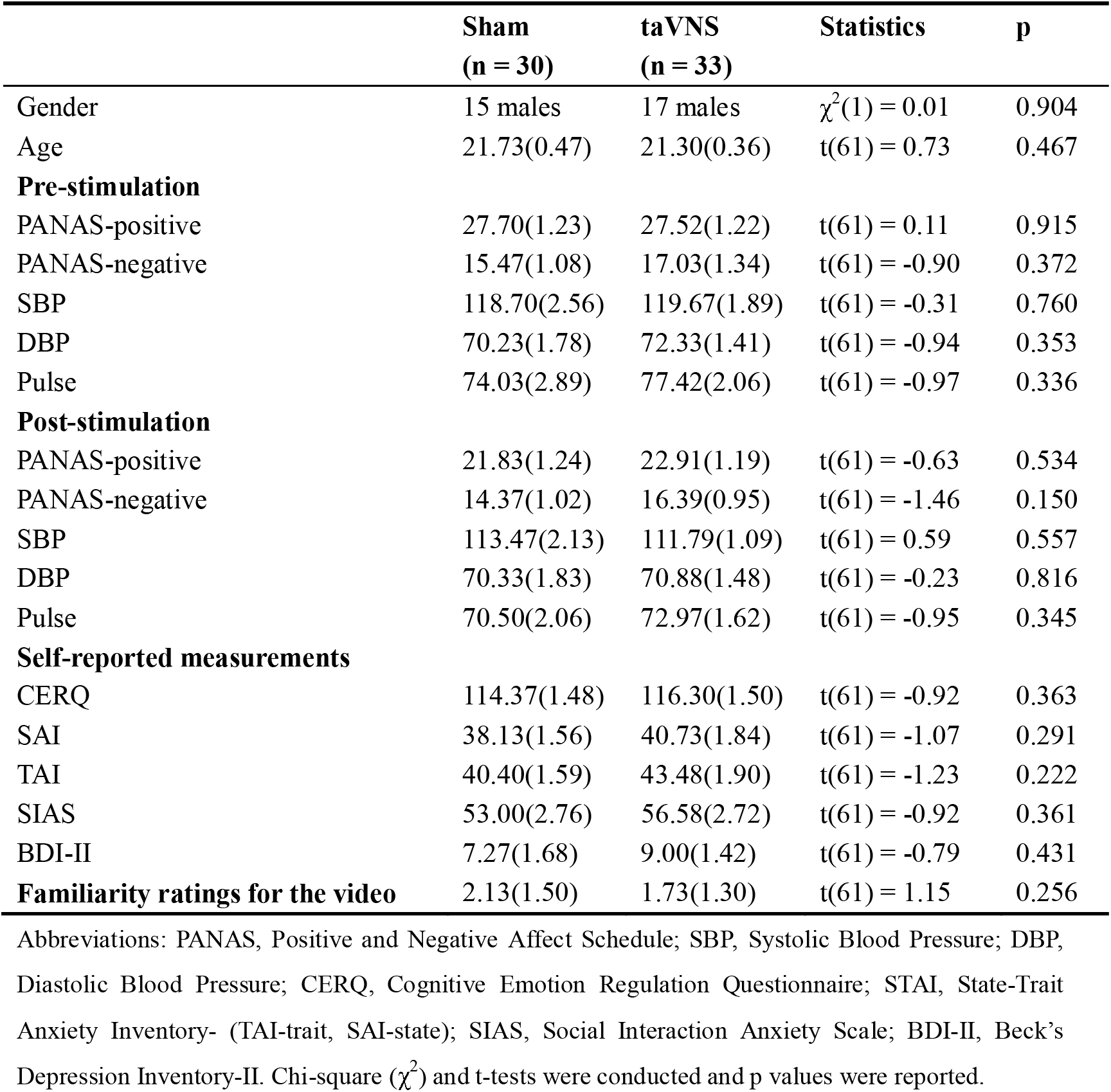
Demographic characteristics and self-reported measurements (mean ± SEM).

### Cross-modal validation of stimulus-evoked activity

To validate fNIRS signal reliability during fear video viewing, we compared spatial activation patterns between fNIRS HbO concentrations under sham condition and fMRI BOLD signals. Pearson correlation analysis revealed significant positive associations between fNIRS and fMRI signals across all ROIs (rs > 0.12, FDR-corrected p < 0.05) across 300 time points, demonstrating strong cross-modal spatial coherence (**Figure 2A**). Furthermore, this coherence was most pronounced for the anatomically relevant region. Specifically, the fNIRS-derived signals showed the strongest correlations with fMRI signals from the frontal lobe, compared to the parietal, occipital, and temporal lobes (**Figure S1B**). These results confirmed that during fear processing, fNIRS reliably captures cortical activation patterns in prefrontal regions comparable to those observed with fMRI, indicating robust cross-modal validation of stimulus-evoked activity.

**Figure 2.**
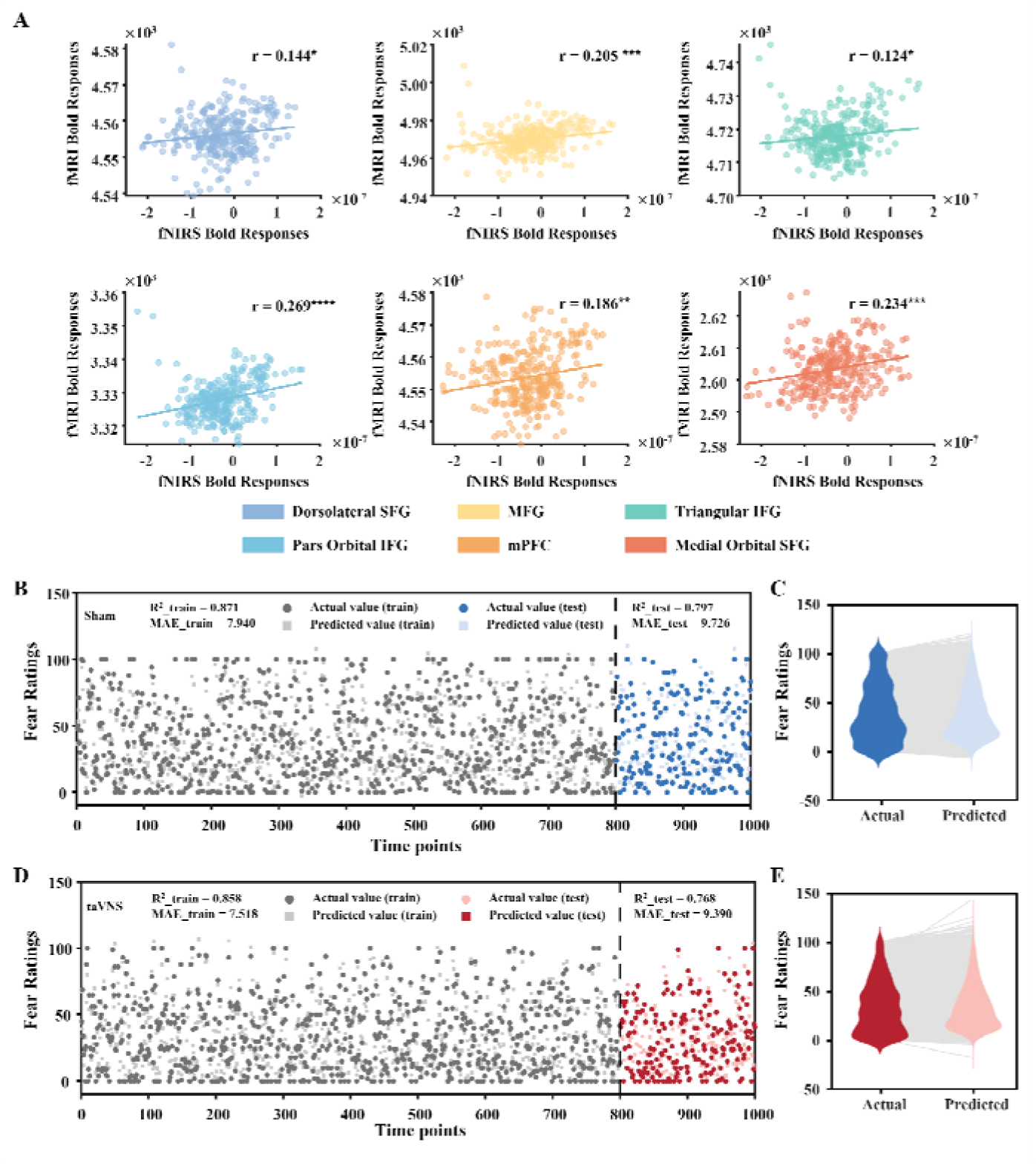
Cross-modal validation of stimulus-evoked activity and fNIRS-based decoding of continuous fear ratings. A. Spatial consistency using Pearson’s correlation between fNIRS and fMRI activation patterns during fear video viewing, demonstrating convergent neural signatures across imaging modalities. B/D. Based on fNIRS signals from the prefrontal cortex, the convolutional neural network (CNN) was employed to predict subjects’ subjective fear ratings. Figures B and D respectively illustrate the performance of one CNN model from each group in decoding subjective fear intensity on the training and test sets; 800 and 200 data points were selected from the training and test sets, respectively, for result visualization. R-square (R^2^) and mean absolute error (MAE) are reported to assess the predictive validity of the model. C/E. Consistency between model-decoded fear intensity (light color) and self-reported fear intensity (dark color) across all sampling points in the test set for the two groups of subjects.

### fNIRS-based decoding of continuous subjective fear ratings

To quantify the relationship between prefrontal cortical activation and subjective fear ratings, we conducted a CNN model mapping 15-channel fNIRS signals to continuous fear ratings. The model demonstrated strong predictive validity across 100 independent cross-validation iterations in both groups (sham: R^2^_train = 0.871±0.013, R^2^_test = 0.797±0.014, MAE_train = 7.940±0.416, MAE_test = 9.726±0.379; taVNS: R^2^_train = 0.858±0.012, R^2^_test = 0.768±0.015, MAE_train = 7.518±0.335, MAE_test = 9.390±0.313). As shown in **Figure 2B**, the CNN successfully reconstructed fear rating dynamics from neural activity patterns. These results demonstrated that prefrontal fNIRS signals robustly encoded continuous subjective fear intensity across stimulation conditions and fNIRS provides a reliable neuroimaging modality for quantitative fear evaluation.

### Context-domain modulatory effects of taVNS on fear responses

In general, there was no differences for subjective fear ratings averaged across the whole task between taVNS and sham group (Sham:38.81±17.18; tavns:33.11±16.03, t = 1.36, p = 0.178). Importantly, time-course analysis identified significant between-group differences in fear ratings during four critical periods (T1-T4, cumulative duration = 50 s) of the 600s fear video, although T3 did not survive permutation-based cluster correction (q_perm_=0.09, **Figure 3A-B**). After adding the fear ratings from an independent sample of 20 participants as a covariate, the results remained consistent. Notably, AI-based moonshot-v1 scene classification characterized each phase as: T1: initial threat exposure (ghosts wandering, lasted 8.67 s, M_sham_±SD =53.71±23.99; M_taVNS_±SD = 35.1±21.86); T2: active danger avoidance (protagonist fleeing, lasted 23.97 s, M_sham_±SD =42.60±27.78; M_taVNS_±SD =29.77±20.66); T3 as an exploratory finding: anticipatory dread (imminent ghost appearance, lasted 9.69 s, M_sham_±SD =51.37±29.59; M_taVNS_±SD = 38.63±27.59); T4: emotional resolution (love suicide denouement, lasted 7.65 s, M_sham_±SD =18.39±22.64; M_taVNS_±SD = 9.21±12.28). The remaining periods with high and moderate fear ratings were characterized by expressions of fear, or physical signs of pain and suffering, together providing quantitative control for fear content rather than fear ratings alone that may drive these localized effects of taVNS. The fear ratings were significantly lower during T4 compared to all other time windows (T1 vs. T4, p < 0.001; T2 vs. T4, p = 0.002; T3 vs. T4, p < 0.001), with no significant differences observed among T1, T2, and T3 (all ps > 0.08) in sham condition. Notably, while fear ratings differed between groups, SCR showed no significant variations (all ps > 0.19), indicating taVNS specifically modulates fear perception rather than generalized physiological arousal.

**Figure 3.**
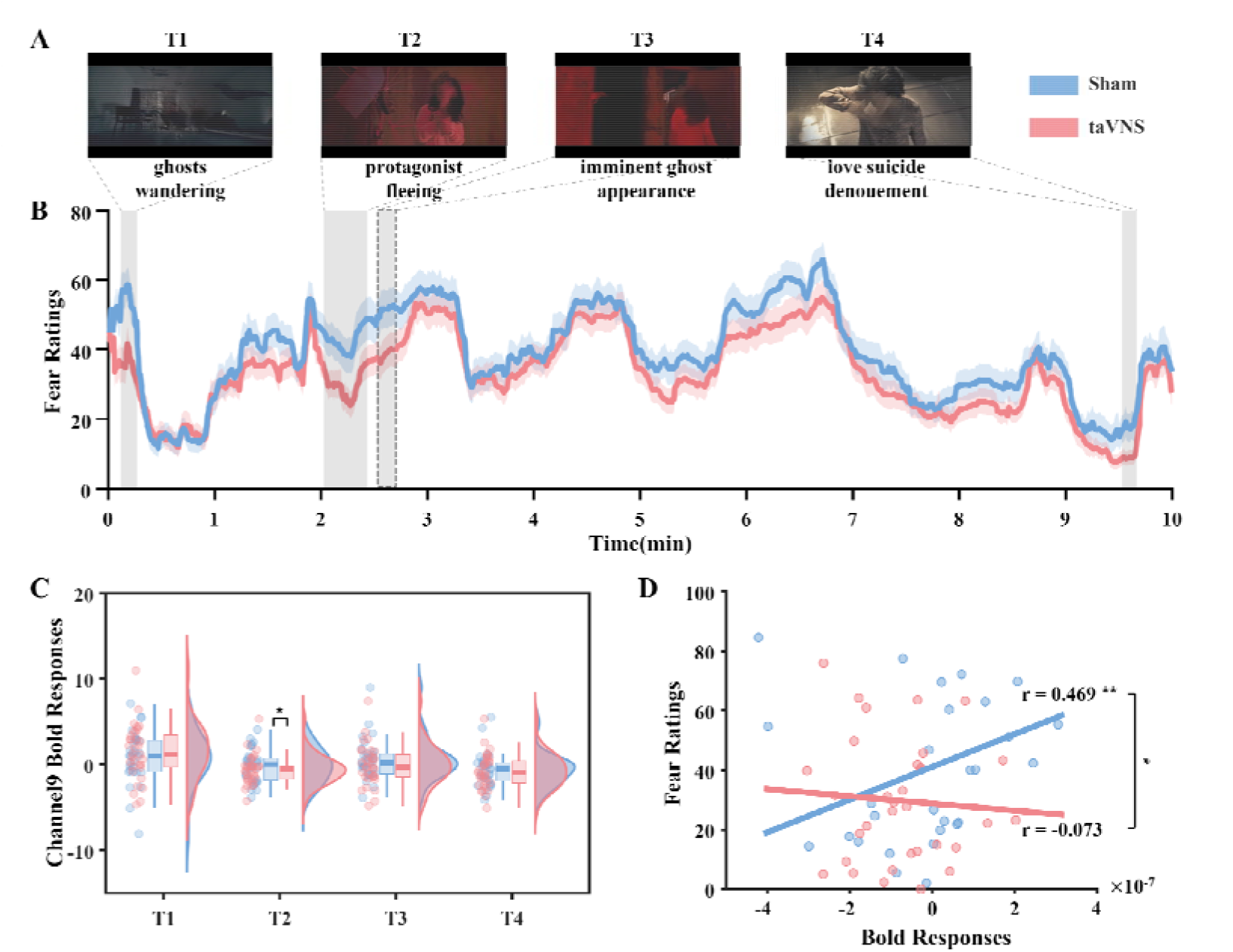
taVNS modulation of fear responses and associated neural correlates. A. Four critical fear periods (T1-T4) were characterized by AI-based moonshot-v1 scene classification. B. taVNS diminished subjective fear during four critical fear periods using t-tests with permutation-based cluster correction. The effect at T3 did not survive correction (q_perm_=0.09) with dashed line. The shaded area represents the standard deviation. C. taVNS decreased neural activation in Channel 9 especially during T2 with permutation correction. *p<0.05 with permutation test. D. The associations using Pearson correlation between subjective fear rating and activation in Channel 9 during T2 in the two groups. Differences in coupling patterns between groups was assessed by Fisher’s z. *p<0.05.

### taVNS specifically decreased mPFC activation and associations with anxiety

During the critical threat-avoidance period (T2), we observed significantly reduced activation in channel 9 (mPFC, MNI coordinates −13, 68, 24, see Table S1& S2) in the taVNS group but not the sham controls (t=2.181, Cohen’s d=0.380, permutation p=0.029; **Figure 3C**). Neural-behavioral correlation analysis demonstrated a significant positive association between fear ratings and mPFC activation in sham controls (r=0.469, p=0.009), which was completely abolished in the taVNS group (r=-0.073, p=0.688), with statistically distinct coupling patterns between groups (Fisher’s z=2.19, p=0.029, **Figure 3D**). These results indicated that taVNS may attenuate fear responses through selective inhibition of mPFC hyperactivation, effectively decoupling this key regulatory region from threat appraisal circuitry. Notably, a significant moderation effect (R^2^=0.22, F=5.62, p =0.002) was found that the interaction between activity of mPFC and state anxiety significantly predicted fear ratings during T2 in both groups (B= −0.388, SE =0.16, t =-2.47, p =0.017). Thus, state anxiety significantly moderated the associations between the activity of mPFC and fear ratings, with post hoc tests using the Johnson-Neyman approach indicating that this moderation was driven by a selective positive association between mPFC activity and fear ratings in participants with lower state anxiety (t = 4.01, p = 0.0002, **Figure 4D**). Further, we applied a data-driven Hidden Markov Modeling (HMM) to obtain group-level estimates of dynamic brain states elicited by the video stimuli. The results revealed that, compared to the sham group, the taVNS group exhibited a significant decrease in fractional occupancy in State 2. This state was characterized by broadly higher activation intensity distributed across mPFC channels (channels 6–10) relative to the other four states (see Supplementary), suggesting that mPFC serves as a core prefrontal configuration robustly recruited to process and regulate fear response by taVNS.

**Figure 4.**
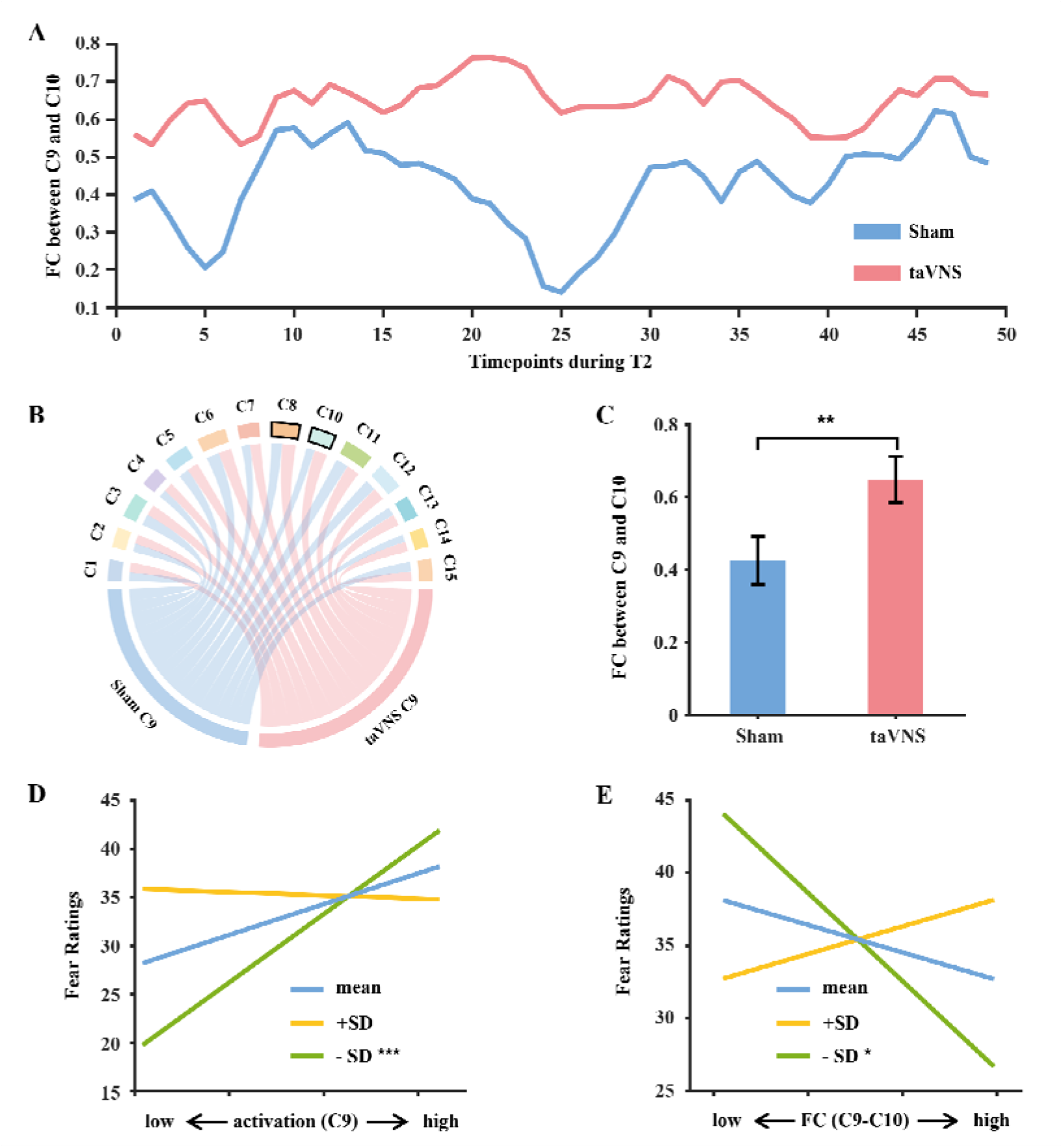
taVNS modulation of mPFC functional connectivity and anxiety-dependent effects. A. Group differences between sham and taVNS on functional connectivity using dynamic conditional correlation between C9 and C10. B. Functional connectivity between C9 and the other channels in the two groups during T2. Black boxes represent channels with significant differences in functional connectivity. C. Group comparation on functional connectivity between C9 and C10 during T2.** p_perm_<0.01 D. Moderation effects of state anxiety on the associations between the activity of mPFC and fear ratings. E. Moderation effects of state anxiety on the associations between the mPFC intra-connectivity and fear ratings.

### taVNS enhanced local network integration and associations with anxiety

Dynamic conditional correlation analysis revealed significantly strengthened functional connectivity within the mPFC network during threat avoidance (T2 period) in the taVNS group compared to sham controls. Specifically, we observed increased Channel 9-Channel 8 (t= 2.21, Cohen’s d=-0.554, permutation p=0.032, **Figure S3**) and Channel 9-Channel 10 (t= 2.93, Cohen’s d=-0.731, permutation p=0.005, **Figure 4A-C**) connectivity. These coordinated enhancements in local network integration suggest that taVNS promotes adaptive and tempered subjective fear through strengthened intra-mPFC communication.

We observed a marginal moderation effect of state anxiety on the mPFC intra-connectivity and fear rating relationship during the escape sequence (T2) (interaction R^2^=0.12, F=2.63, p =0.058). The anxiety × mPFC intra-connectivity interaction significantly predicted fear ratings across both groups (B= 2.09, SE =0.92, t =2.27, p =0.027). Post hoc analysis using the Johnson-Neyman technique revealed this moderation was driven by a selective positive association between mPFC intra-connectivity and fear ratings in participants with lower state anxiety (t=2.63, p=0.011; **Figure 4E**).

## Discussion

This study investigated the effects of non-invasive taVNS on continuous fear evaluation in a naturalistic dynamic context. Our findings demonstrated that: (1) fNIRS-derived prefrontal signals correlated with fMRI measurements, and a CNN model confirmed that prefrontal features predicted instantaneous fear ratings in both groups; (2) taVNS reduced fear responses during specific threat-related time windows, accompanied by decreased mPFC activity and enhanced functional integration within the mPFC network; and (3) these neural changes were moderated by state anxiety levels. These preliminary results suggest that taVNS can acutely modulate fear-related neural and behavioral responses. This finding motivates future studies to test whether similar effects extend to extinction-learning paradigms and clinical outcomes in fear-related disorders and, specifically, whether it may serve as an early adjunctive intervention to facilitate initial fear reduction during exposure therapy [35].

In our study, we found prefrontal activity predicted instantaneous fear ratings during video viewing in both stimulation groups. This supports the key role of prefrontal regions in subjective fear evaluation, in line with previous results that prefrontal regions are not essential for fear learning [7] but are involved in fear extinction [10, 11]. As a cross-modal validation of stimulus-evoked cortical signals, we observed high spatial coherence between fNIRS-derived HbO patterns (under sham) and fMRI BOLD patterns during viewing of the same naturalistic fear video. This correspondence supports the fidelity of the fNIRS measurements for capturing prefrontal activation during naturalistic fear processing, consistent with prior work demonstrating broad comparability between fNIRS and fMRI for cortical functional integration across spatial scales in both resting-state [61] and task contexts [62]. While we observed no significant sex differences in fNIRS-based fear ratings or ROI activity, this factor is unlikely to limit the generalizability of our cross-modal correspondence to the fMRI dataset. Future validation using sex-balanced multimodal samples remains necessary to confirm generalizability across sexes.

Importantly, fMRI evidence confirms that cymba conchae stimulation activates central vagal projections, including the nucleus of the solitary tract, spinal trigeminal nucleus, locus coeruleus, and amygdala [20], as well as the prefrontal cortex during task-free scanning [63]. Building on this foundation, our study employed AI-based moonshot-v1 content analysis to identify that taVNS (vs. sham) selectively suppressed mPFC activity during four fear-related epochs characterized by: (1) ghost encounters, (2) active danger avoidance, (3) anticipatory dread, and (4) suicide denouement. This finding carries particular significance given the mPFC’s established role in fear expression. Prior work shows that mPFC exhibited a sophisticated temporal coding mechanism with phase-specific inhibition bidirectionally controlling fear responses (ascending phase inhibition blocks fear, descending phase inhibition potentiates it) [64, 65]. This establishes a precise phase-locked pathway by which mPFC dynamically coordinates neural assemblies to modulate fear responses. Notably, in the sham condition, mPFC activity showed sustained positive associations (>20s) with fear ratings during active danger avoidance periods, an association abolished by taVNS. These findings position taVNS as a potential non-invasive method for implementing domain-specific mPFC modulation, effectively serving as a negative emotion regulation strategy through selective mPFC inhibition.

Exposure-based interventions represent one of the most effective treatments for anxiety disorders [66]. To enhance therapeutic outcomes, researchers have proposed that combining evidence-based interventions may yield greater benefits than monotherapy, a premise supported by studies augmenting exposure therapy with pharmacological agents such as oxytocin [41] or losartan [12]. Beyond pharmacotherapy, neuromodulation approaches like taVNS may offer a promising method for real-time modulation of fear-related neural processing. Our findings demonstrate that taVNS consistently suppressed mPFC activity across key fear-processing phases (ghost encounters, active danger avoidance, anticipatory dread, and suicide denouement), with particularly pronounced effects during threat detection and avoidance behaviors. This suggests that taVNS may attenuate maladaptive defensive responses by regulating prefrontal hyperreactivity, a mechanism that could enhance exposure therapy efficacy by facilitating early fear reduction and improving treatment engagement.

Notably, taVNS-enhanced mPFC connectivity during active danger avoidance suggesting improved local prefrontal network coordination that may facilitate more efficient threat appraisal. This finding aligns with recent evidence demonstrating that taVNS-induced cognitive improvements correlate with increased mPFC functional connectivity [67]. The temporal dynamics of such connectivity changes appear particularly relevant - healthy controls show progressive mPFC connectivity increases during fear extinction learning, a pattern notably absent in anxiety and PTSD patients [68]. Importantly, these connectivity impairments significantly correlate with clinical symptom severity, highlighting their pathological relevance [68]. Moreover, the degree of connectivity enhancement during extinction learning predicts the strength of extinction memory recall after 24 hours [69]. Collectively, this work raises the possibility that taVNS-induced mPFC connectivity modulation could support neural processes involved in subjective fear adaptation. Additionally, the associations between brain alterations (i.e., mPFC activity or within-mPFC integration) and continuous fear ratings were moderated by anxiety state, reflecting a synergistic relationship between anxiety and fear. Specifically, persistent anxiety potentiates fear responses upon threat exposure [70].

The SCR has previously been validated as a measure of autonomic arousal in traditional Pavlovian conditioning paradigms [8] and predator exposure protocols [71]. Notably, recent studies have employed advanced neural decoding approaches to demonstrate that this hard-wired physiological arousal response – as assessed by SCR - is represented in neural representations that are distinguishable from the neural representations of the subjective and conscious affective experience[14–16]. Despite detecting changes in subjective fear ratings and neural activity/connectivity, the absence of SCR differences could reflect several possibilities: (1) taVNS may primarily modulate the subjective aspect of fear rather than the automatic arousal; (2) the naturalistic viewing paradigm with continuous rating demands reduced the sensitivity of SCR to group differences, or (3) a small physiological effect exists that our study was underpowered to detect. Regardless, the behavioral and neural findings should be interpreted with the understanding that they were not accompanied by significant changes in peripheral physiology.

Several limitations of this study should be acknowledged. First, our investigation focused exclusively on six prefrontal cortical areas using fNIRS, given the established role of the mPFC in fear processing. This methodology, however, cannot assess deeper subcortical and limbic regions (e.g., amygdala, hippocampus, brainstem nuclei, insula) that are essential to fear circuitry [71] and engaged by taVNS [19–21]. Therefore, future combined fMRI-taVNS studies are needed to characterize these regions and the full network dynamics. Second, group differences were confined to specific, threat-related time windows, rather than periods with higher fear ratings. Together with the quantitative evidences of fear ratings from an independent sample, this finding indicates that taVNS effects are context-dependent and may be diluted by averaging across an entire task. Future studies should therefore incorporate detailed stimulus annotation and employ multiple naturalistic paradigms to assess the robustness and generalizability of these effects. Third, regarding the CNN predictive modeling, it is important to note that our validation strategy employed a random split of pooled data from all participants. While we have mitigated the model’s ability to learn sequential dependencies in the data to a certain extent by means of global random shuffling, the presence of data from the same participants in both training (80%) and testing sets (20%) means the current results reflect the model’s ability to decode continuous fear states within the studied cohort. Future studies employing participant-level cross-validation are considered to determine whether these neural signatures are fully generalizable and to rule out potential participant-specific confounds. Finally, the present study tested only acute stimulation effects in healthy volunteers during naturalistic viewing, without extinction-learning paradigms, clinical symptom measures, or longitudinal follow-up. Despite this, the observed modulation of brain-fear coupling by state anxiety indicates potential translational relevance. Systematic clinical studies are needed to establish whether these effects generalize to anxiety disorders and to treatment-relevant learning processes. A null finding that we did not observe group differences in SCR weakens the claim that taVNS modulated fear processing generally. Future studies should combine autonomic, neural, and subjective measures to clarify which components of fear are most amenable to taVNS modulation. Key parameters requiring further investigation include optimal stimulation timing (pre-, during, or post-extinction training) and individual variability in treatment response. Future studies should also implement a fully double-blind design (i.e. dual-electrode wearing + algorithmic random activation) to further eliminate experimenter bias and potential expectancy effects, thereby improving internal validity.

Our findings provide preliminary evidence that acute taVNS is associated with context-dependent reductions in subjective fear during naturalistic viewing, accompanied by alterations in prefrontal dynamics including decreased mPFC activity and increased within-mPFC functional integration. Collectively, these findings support the potential of taVNS as a promising early adjunct to exposure-based therapies for fear-related disorders (e.g. anxiety disorder), warranting further clinical investigation.

## Data Availability

All data produced in the present study are available upon reasonable request to the authors.

## Availability of Data and Materials

Data in the present study can be made available upon request to the primary contact author, and code are available at https://github.com/zhaolab205/taVNS_effects.

