## Supplementary Information for "Transcutaneous auricular vagus nerve stimulation regulates subjective fear in naturalistic contexts via modulation of prefrontal neural dynamics"

**Supplementary Methods:**

**Stimulus intensity adjustment procedure**

During the calibration procedure, participants received increasing and decreasing series of stimulation trials, and reported their subjective sensation of the stimulation on a 7-point Likert scale (1: feeling nothing, to 7: painful). The increasing series of trials started from 0 mA and elevated in steps of 0.1 mA until participants reported a “painful” sensation of 7. On the other hand, in decreasing series of trials, the “painful” intensity was repeated and then reduced in steps of 0.1 mA until a subjective sensation of “feeling nothing” was experienced. This procedure was performed twice and the final stimulation intensity for each participant was calculated based on the average of four intensity values (i.e., two from increasing and two from decreasing trials) that were rated as 5 (i.e., obvious “tingling” sensation but not painful).

**MRI data acquisition and preprocessing**

Functional MRI data were acquired using a T2*-weighted echo-planar imaging (EPI) sequence with the following parameters: repetition time (TR) = 2000 ms, echo time = 30 ms, 36 slices, slice thickness = 3.8 mm, field of view = 200 × 200 mm, resolution = 64 × 64, flip angle = 90°, 3.125 × 3.125 × 3.8 mm voxels. High-resolution T1-weighted images were acquired using a 3D spoiled gradient recalled (SPGR) sequence (176 slices, repetition time = 8.22 ms, echo time = 3.15 ms, field of view = 256 × 256 mm, resolution = 256 × 256, flip angle = 8°, 1 × 1 × 1 mm voxels).

The MRI preprocessing pipeline included: (1) spatial normalization to the ICBM 152 Nonlinear Asymmetrical template version 2009c [1, 2] was performed through nonlinear registration with the antsRegistration tool of ANTs v2.3.3 [3], using brain-extracted versions of both T1w volume and template; (2) brain tissue segmentation of cerebrospinal fluid (CSF), white matter (WM) and gray matter (GM) was performed on the brain-extracted T1w using FAST (FSL v6.0.5.1) [4]; (3) five initial volumes of fMRI data were removed to allow for image intensity stabilization; (4) functional data was slice time corrected using 3dTshift from AFNI [5] and motion corrected using mcflirt (FSL v6.0.5.1) [6]; (5) co-registration to the corresponding T1w using boundary-based registration [7] with six degrees of freedom, using FLIRT (FSL) [8]; (6) motion correcting transformations; (7) BOLD-to-T1w transformation and (8) T1w-to-template (MNI) warp (interpolated to 2 mm isotropic voxels) were concatenated and applied in a single step using ants ApplyTransforms (ANTs v2.3.3) using Lanczos interpolation. Spatial smoothing was performed using an 8 mm full-width at half-maximum (FWHM) Gaussian kernel.

**Data-driven Hidden Markov Modeling**

The Hidden Markov Modeling (HMM) [9–11] assumes that time-series data can be described using a hidden sequence of a finite number of states. More explicitly, if vector $y_{t}$ represents the data and $s_{t}$ represents the hidden state at time point $t$, we assume that:

$$y_{t}|s_{t}=k \sim Multivariate Gaussian(\mu_{k,}\Sigma_{k})$$

where $\mu_{k}$ is a vector with elements equal to the number of channels, containing the mean blood oxygen level-dependent (BOLD) activation, and $\Sigma_{k}$ is the covariance matrix (number of channels by number of channels) codifying the variances and covariances between channels when state $k$ is active. This is referred to as the observational model, which characterizes the distribution of each state $k$ through parameters ($\mu_{k}, \Sigma_{k}$). We use a multivariate Gaussian distribution here.

Furthermore, the state sequence is regularized by modeling the transition probabilities between all pairs of brain states. That is, before observing the data, the probability,$Pr$,of a given state being active at time point $t$ depends on which state was active at time point $t-1$:

$$\Pr\left( s_{t}=k | s_{t-1}=j \right)=A_{jk}$$

where $A$ refers to the transition probabilities. Within matrix $A$, we can further distinguish between the on-diagonal elements, $A_{kk}$, which control the persistence of each state, and the off-diagonal elements, $A_{jk}$ (with $j\neq k$), which refer to the actual transitions. Finally, a parameter $\eta$ encodes the initial state probabilities for each scanning session. Therefore, according to this formulation, the observed data at each time point are effectively modeled as a mixture of Gaussian distributions, with weights given by $Pr(s_{t}=k)$.

We applied the HMM to the concatenated time-series data of all subjects to obtain group-level estimates of the brain states. We employed an inference algorithm to estimate the following parameters from the observed data: the parameters defining the posterior distribution of each state (i.e., the state mean $\mu_{k}$ and covariance $\Sigma_{k}$), the activation probability of each state at each time point (i.e., the state sequence $s_{t}$), and the transition probabilities between pairs of states ($A_{jk}$).Specifically, working within a Variational Bayes (VB) framework, we imposed additional factorization assumptions on the posterior distribution to achieve an analytical approximation of the model's posterior at a reasonable computational cost. In practice, VB inference typically employs a strategy of alternately updating each group of parameters, iterating among different parameter sets until convergence is reached.

An important byproduct of VB inference is the free energy. Serving as an approximation of the Bayesian model evidence, it can be utilized to evaluate the model's goodness-of-fit to the data and to guide model selection. To determine the optimal number of hidden states, K, we used free energy as the evaluation metric, computing it for K values ranging from 2 to 8, and identified the optimal number of states using the elbow method. For each K, the algorithm was independently run 5 times, and the iteration yielding the optimal free energy was selected. Finally, formal inference was conducted based on the established optimal number of states, repeated 20 times independently, with the iteration achieving the best free energy (i.e., the best fit) selected as the final model.

**Supplementary Results:**

We applied a HMM to obtain group-level estimates of dynamic brain states elicited by the video stimuli. Prior to formal estimation, we determined the optimal number of hidden states (K = 5, **Figure S4C**) using variational free energy combined with the elbow method. The HMM was then executed with K = 5 to derive the final state distribution (**Figure S4A** and **B**). Based on this, we calculated subject-specific temporal metrics, specifically, fractional occupancy (FO). Two-sample t-tests were employed to examine group differences in FO and fear ratings across the five states, with permutation testing applied for rigorous statistical correction.

The results revealed that, compared to the sham group, the taVNS group exhibited a significant decrease in FO in State 2 (t = 2.430, permutation p = 0.020, **Figure S5E**).Spatial activation profiles indicated that State 2 was characterized by broadly higher activation intensity in the mPFC compared to other states (State 2 vs. 1, t = 4.682, p__FDR_ < 0.001; State 2 vs. 3, t = 4.365, p__FDR_ < 0.001; State 2 vs. 4, t = 3.786, p__FDR_ < 0.001; State 2 vs. 5, t = 0.842, p__FDR_ = 0.411; **Figure S5D**), suggesting that the mPFC serves as a core prefrontal configuration robustly recruited by taVNS to process fear response. Concurrently, during the occurrence of State 2, fear ratings in the taVNS group were significantly lower than those in the sham group (t = 2.121, permutation p = 0.036, **Figure S5F**).

**Figure S1** **A**. Correlations among six fNIRS-based prefrontal cortical regions, and **B.** correlations between these six prefrontal fNIRS ROIs and four major cerebral lobes (frontal, parietal, occipital, temporal) from fMRI.

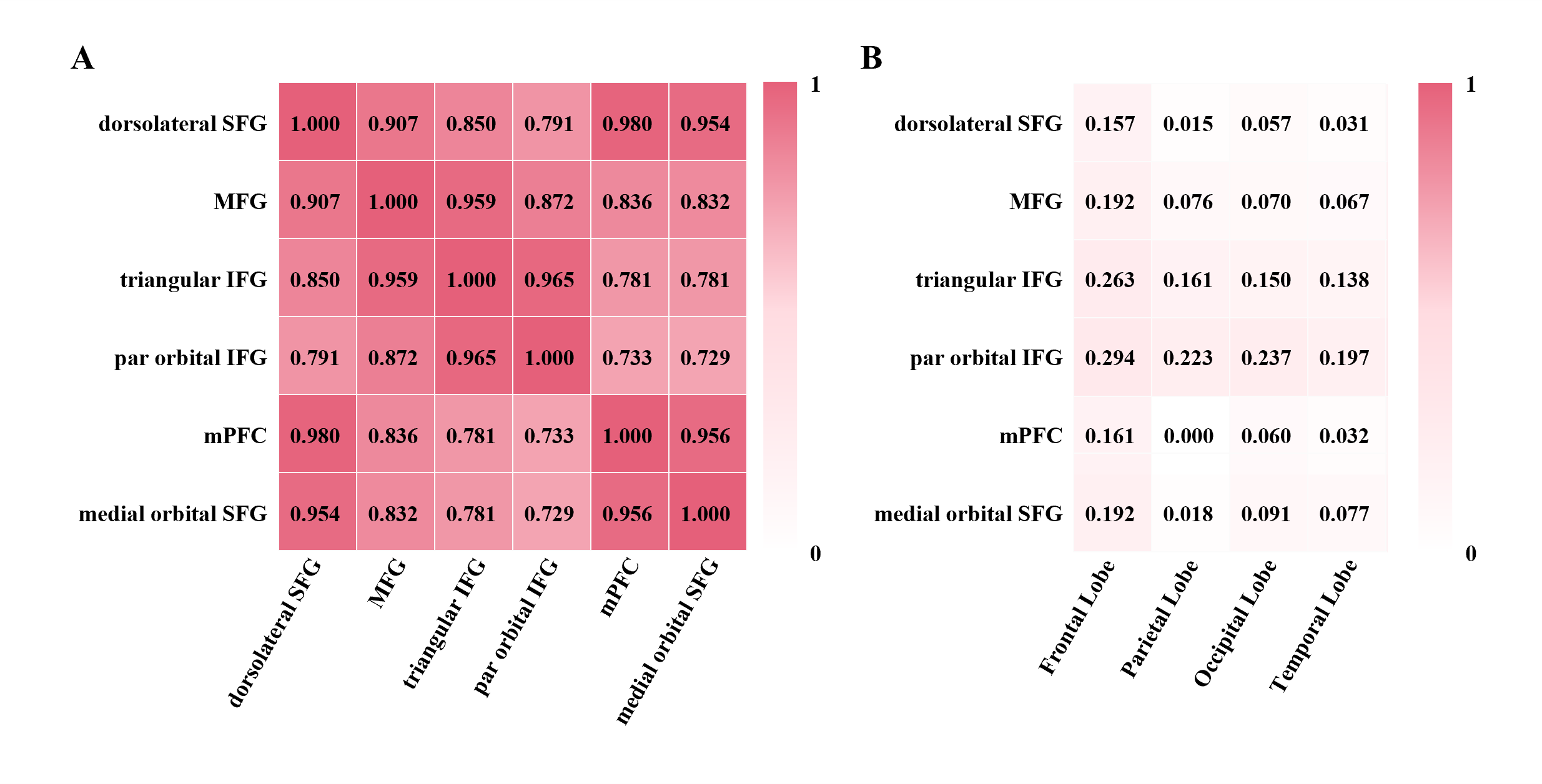

**Figure S2** Predictive validity of the CNN model across two groups at 100–300 epochs.

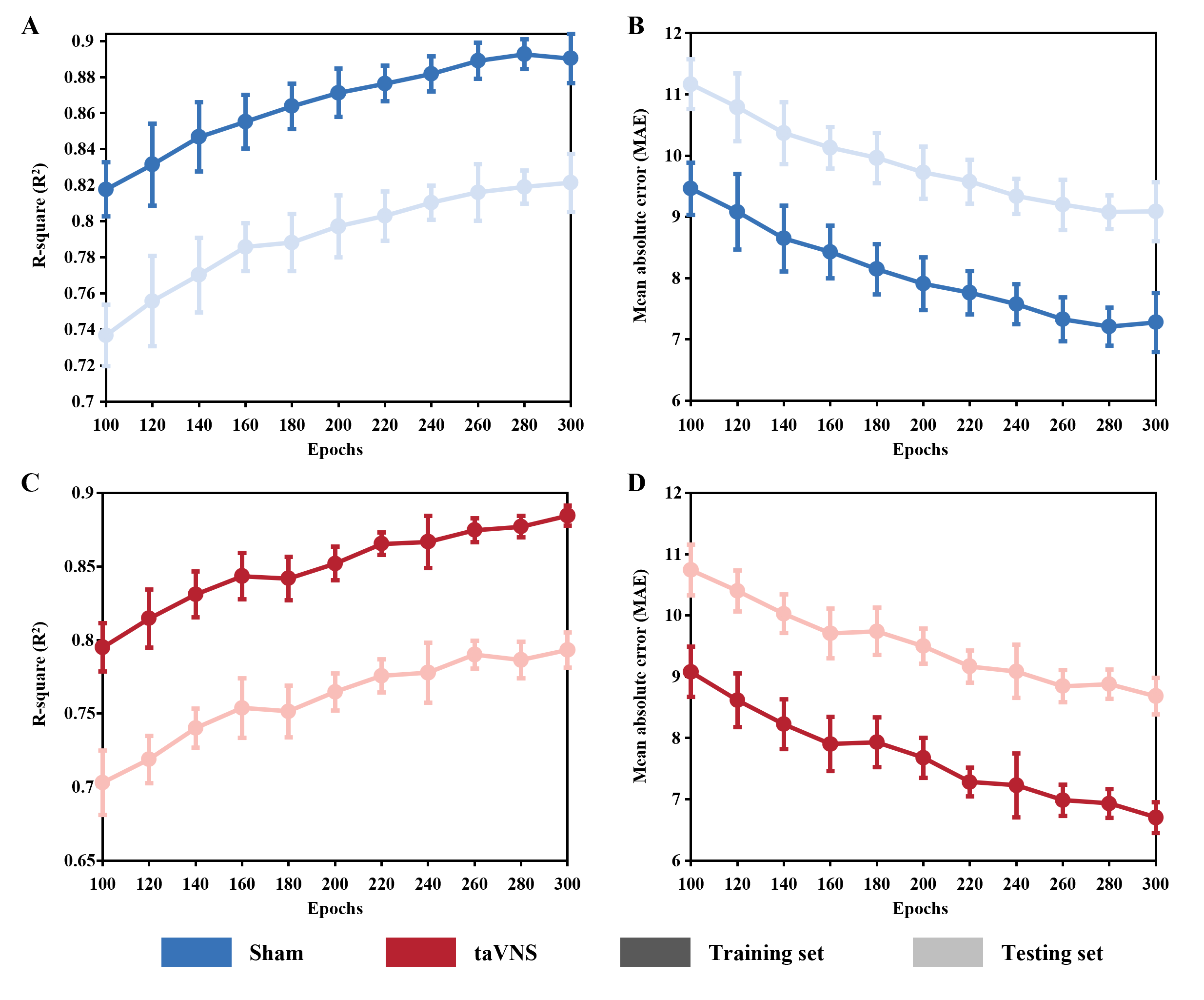

**Figure S3 A.** Group differences between sham and taVNS on dynamic functional connectivity between C9 and C8. **B.** Group comparation on functional connectivity between C9 and C8 during T2.

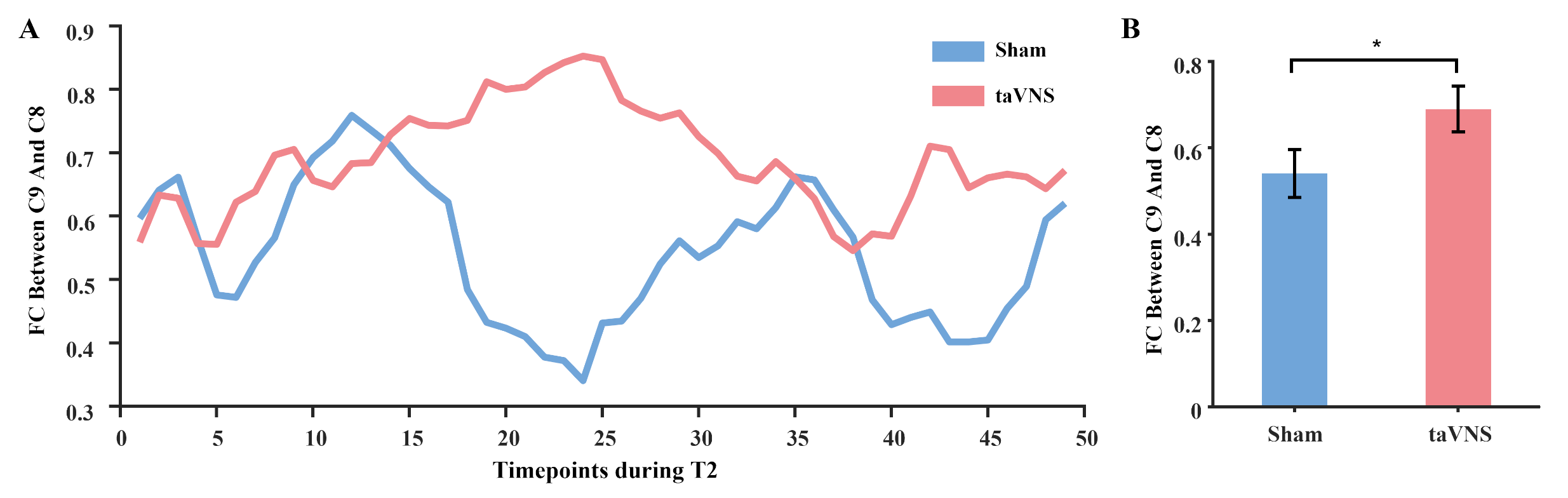

**Figure S4** Results of HMM state analysis for stimulation groups. **A/B.** Occurrence probability of each state at each time point for the sham and taVNS groups. **C.** Optimal number of states. The red circle highlights the optimal number of states determined by the elbow criterion. **D.** Mean activation across 15 channels for each state. **E.** Group differences in FO. **F.** Group differences in subjective fear ratings. * indicates p_perm_ < 0.05.

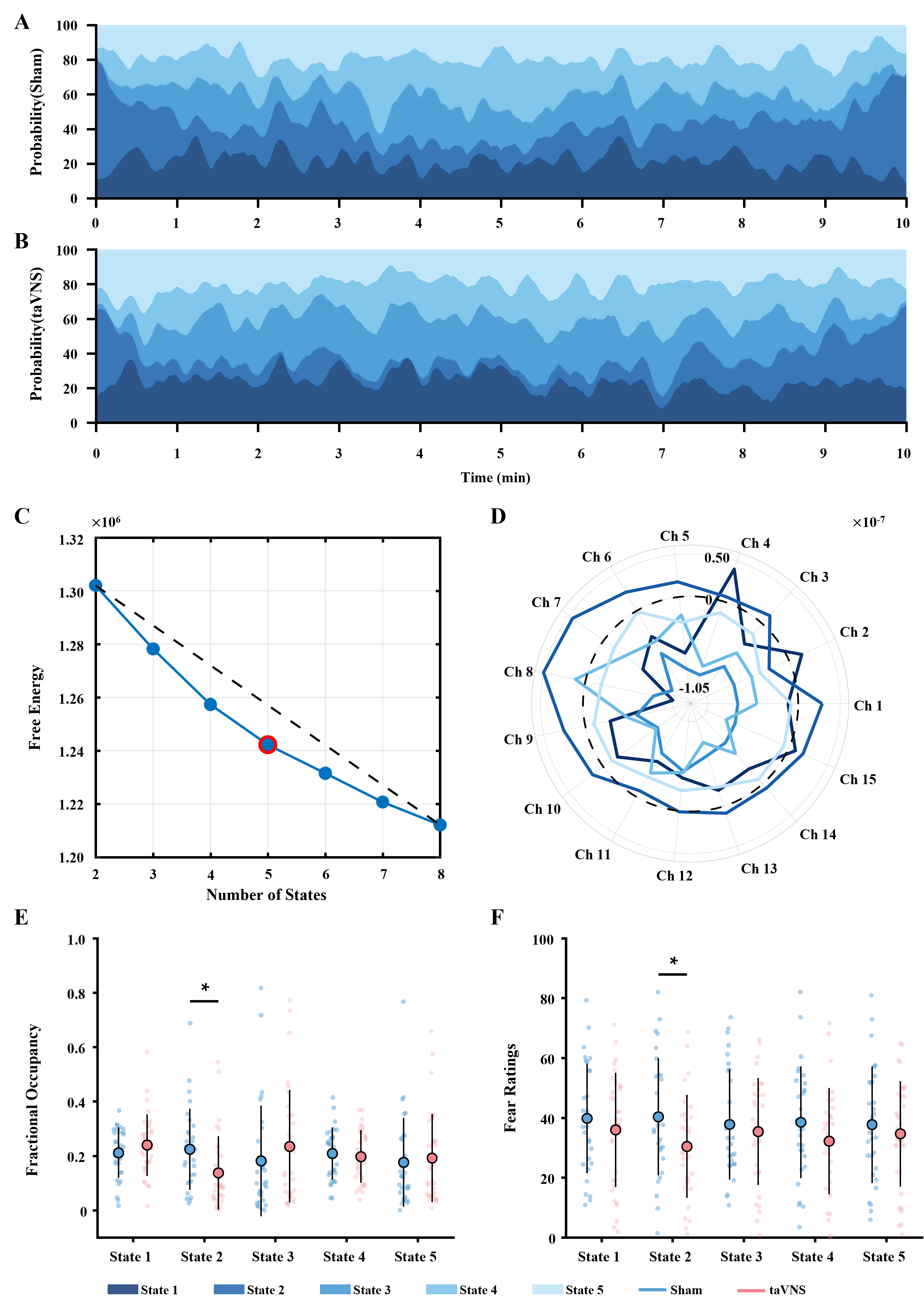

**Table S1** MNI coordinates corresponding to each channel

| **Channel** | **X** | **Y** | **Z** | **Hemisphere** |
| --- | --- | --- | --- | --- |
| Channel 1 | 54.67 | 44.00 | 0.00 | Right |
| Channel 2 | 48.00 | 52.67 | 12.67 | Right |
| Channel 3 | 36.33 | 58.33 | 24.67 | Right |
| Channel 4 | 39.00 | 64.67 | 0.00 | Right |
| Channel 5 | 26.33 | 69.67 | 12.67 | Right |
| Channel 6 | 13.67 | 69.00 | 24.67 | Right |
| Channel 7 | 14.33 | 73.00 | 0.33 | Right |
| Channel 8 | 0.33 | 68.00 | 11.33 | Center |
| Channel 9 | -13.00 | 68.00 | 24.00 | Left |
| Channel 10 | -14.33 | 73.00 | 0.00 | Left |
| Channel 11 | -26.00 | 68.00 | 12.00 | Left |
| Channel 12 | -35.67 | 57.67 | 23.67 | Left |
| Channel 13 | -38.00 | 63.00 | 0.00 | Left |
| Channel 14 | -45.67 | 51.67 | 12.33 | Left |
| Channel 15 | -52.33 | 43.67 | -0.33 | Left |

**Table S2** Channels contained in each brain area

| **Brain region** | **Channel** |
| --- | --- |
| Dorsolateral SFG | CH5 CH6 CH7 CH9 CH10 CH11 |
| MFG | CH2 CH3 CH4 CH12 CH13 CH14 |
| Triangular IFG | CH1 CH2 CH14 CH15 |
| Par Orbital IFG | CH1 CH15 |
| mPFC | CH6 CH7 CH8 CH9 CH10 |
| Medial Orbital SFG | CH7 CH10 |

SFG, superior frontal gyrus; MFG, middle frontal gyrus; IFG, inferior frontal gyrus; mPFC, medial prefrontal cortex.

**Table S3** Subjective ratings for the stimulation adverse effects (mean ± SEM).

|  | **Sham**  **(n = 30)** | **taVNS**  **(n = 33)** | **Statistics** | **p** |
| --- | --- | --- | --- | --- |
| Headache | 2.07(0.23) | 1.61(0.15) | t(61) = 1.71 | 0.093 |
| Nausea | 1.37(0.11) | 1.33(0.14) | t(61) = 0.18 | 0.857 |
| Skin irritation under the electrode | 2.07(0.20) | 2.64(0.23) | t(61) = -1.84 | 0.070 |
| Vigilant | 2.77(0.26) | 3.00(0.25) | t(61) = -0.64 | 0.524 |
| Unpleasant feelings | 2.17(0.22) | 2.09(0.19) | t(61) = 0.26 | 0.793 |
| Dizziness | 1.67(0.16) | 1.52(0.19) | t(61) = 0.61 | 0.543 |
| Neck pain | 1.80(0.23) | 1.39(0.16) | t(61) = 1.50 | 0.140 |
| Muscle contractions in the neck | 2.00(0.23) | 1.64(0.17) | t(61) = 1.31 | 0.194 |
| Stinging sensation in the ear | 3.03(0.26) | 3.42(0.27) | t(61) = -1.05 | 0.298 |
